# Validity of a deep learning algorithm for detecting wheezes and crackles from lung sound recordings in adults

**DOI:** 10.1101/2022.11.18.22282442

**Authors:** Hasse Melbye, Johan Ravn, Mikolaj Pabiszczak, Lars Ailo Bongo, Juan Carlos Aviles Solis

**Affiliations:** General Practice Research Unit, UiT The Arctic University of Norway; Medsensio AS, Tromsø, Norway; Department of Computer Science, UiT The Arctic University of Norway

**Keywords:** Deep learning, algorithms, crackles, wheezes

## Abstract

We validated our state-of-the-art deep learning algorithm for detection of wheezes and crackles in sound files by comparing the classification of our algorithm with those of human experts. We had two validation sets classified by experienced raters that were not used to train the algorithm with 615 (A) and 120 (B) sound files, respectively. We calculated Area Under Curve (AUC) of the algorithm’s probability scores for wheezes and crackles. We dichotomized the scores and calculated sensitivity and specificity as well as kappa agreement. In set A, the AUC was 0.88 (95% CI 0.84 – 0.92) for wheezes and 0.88 (95% CI 0.84 – 0.92) for crackles. The sensitivities and specificities of the labels were 81% and 89% for wheezes and 67% and 96% for crackles. In set B, the kappa agreement between the algorithm and the validation set was 0.78 (95% CI 0.58 – 0.99) for wheezes and 0.75 (95% CI 0.59 – 0.92) for crackles. The 24 observers who had rated the same 120 sound files agreed less with the reference classification with a mean kappa of 0.68 for wheezes and 0.55 for crackles. We found the algorithm to be superior to doctors in detecting wheezes and crackles in lung sound files.

## Introduction

Two types of abnormal lung sounds, wheezes and crackles, are commonly found in patients with chronic obstructive pulmonary disease ^1,2^, heart failure ^3^ pneumonia ^4^, and pulmonary fibrosis ^5^, and are regarded to be useful in the diagnosis of these diseases. Their identification during chest auscultation with a traditional stethoscope is hampered by subjectivity and interobserver variation ^6,7^ This problem can be alleviated by letting computers classify sounds recorded by electronic stethoscopes ^8^. In recent years machine learning based algorithms for detecting adventitious lung sound, mainly wheezes and crackles, have been developed and evaluated ^9-12^, and discussions on opportunities and pitfalls are ongoing ^12,13^. There has also been attempts to go one step further and use lung sounds for a direct diagnosis of lung diseases ^14-18^. In this study we focus on automatic identification of wheezes and crackles and we have validated our state-of-the-art deep learning algorithm for detecting wheezes and crackles trained by recordings from a general population. We have evaluated the algorithm against human ratings of wheezes and crackles in two validation sets with sound files not used in the training of the algorithm.

## Methods

### The lung sound recordings

The 24198 lung sound files used for training of the algorithm were recorded in 4033 participants of the 7^th^ Tromsø Study. This population-based health survey was carried out between May 2015 and October 2016, and methods and study design have previously been described ^19,20^. All Tromsø residents 40 years and older (n=32 591) were invited to participate and a random sample was selected for a second visit where lung sound recording was included. Lung sounds were recorded at six locations of the chest (Fig 1), 15 seconds at each site, with a Sennheiser microphone inserted in the tube of a Littmann Classic II stethoscope. No preprocessing or filtering was done. Ahead of training the algorithm, the lung sound recordings of the training set were classified in terms of presence of wheezes and crackles,^19^

**Figure 1.**
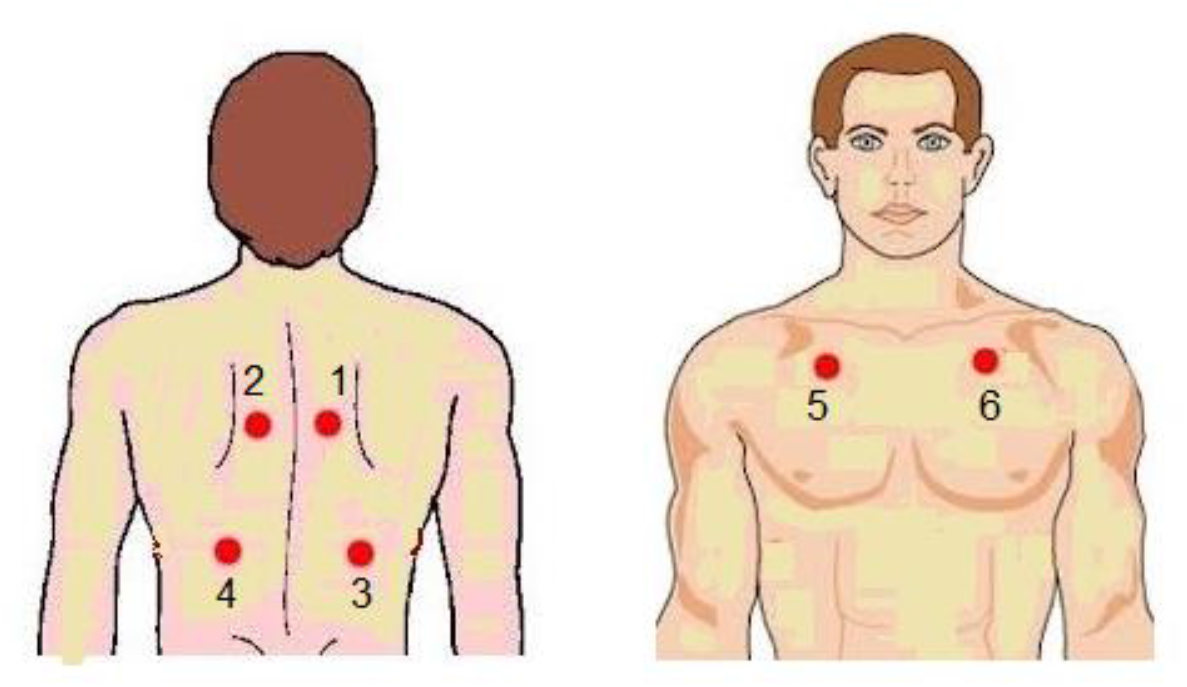
The six recording sites used

In the Tromsø Study lung sounds were also recorded in an additional 2015 participants. These 12090 sound files were not classified and were kept outside the training set. Validation set A is based on these 12090 sound files.

Validation set B consists of the sound files used an interobserver study, in which four lung sound researchers, 20 medical doctors and four medical students took part ^21^.

The lung sounds were recorded in 20 volunteers aged 40 years or more, most of them patients at a rehabilitation center. The same six chest locations as in set A were used, and the validation set consisted of 120 files in total. The sound files had originally a duration of 10 -15 seconds, but had been shortened to avoid sections with noise.

The 7^th^ Tromsø Study was approved by the Regional Committee for Medical and Health Research Ethics. The interobserver study was presented for the Regional Committee for Medical and Health Research Ethics, but the committee waived to evaluate the project since it was considered to be outside the remit of the Act on Medical and Health Research and a written consent was not deemed necessary. No personal information was registered that could link the collected data to the individual subjects.

### Algorithm development

#### Classification of the training set

Presence of wheezes and crackles during inspiration and expiration was determined through a rigorous classification process^19^. At first, two observers (clinicians) independently classified the recordings blinded for other information about the participants. When the observers disagreed, they discussed the actual recordings with a third observer. The recordings judged to contain certain or likely crackles or wheezes, were rated in a second round, where experienced lung sound researchers were among the observers. When listening to and classifying the lung sounds, the observers watched spectrograms of the recordings generated by Adobe Audition© software.

Based on the observers’ ratings, a final decision was made on whether wheezes or crackles were present or not ^19^, which we used as the reference classification of the training set.

#### Training of the algorithm

An architecture based on inception V3 was selected to build the deep learning algorithm^22^. The raw audio data was converted to mel spectrograms, thus turning the audio classification task into an image classification task.

For a single raw audio signal three such spectrograms were extracted, each using different size of the Fourier transform window. Subsequently, the three spectrograms were stacked together forming an RGB format-like image that was used as an input to the machine learning models. The last layer of the architecture is a *sigmoid* activation function that predicts each class (wheezes, crackles or bad quality) with a probability of between 0 and 1. As this is partially a multi-label classification task, a sample can be both wheeze and crackle at the same time (although it cannot be wheeze and bad quality at the same time), we use binary cross-entropy as the loss function for the model.

The models were trained in the 5-fold cross validation procedure. In each fold a model was trained on the fold-specific training set and evaluated on the fold-specific validation set. For each fold, the model that obtained the highest ROC AUC (calculated on the fold-specific validation dataset) was selected as the result model. After selecting the best model in each fold, the next step was to dichotomize the probability scores and select thresholds for each label (wheezes or crackles).

Thresholds are used for deciding whether the probability for a given label is high enough, so that the given label should be assigned (encoded). Distinct sets of thresholds are selected for each of 5 models and the selection of thresholds is performed with the use of the fold-specific validation set (the selection is performed in such a way as to maximize label-specific F1-score). The result model from each of the folds was used to form an ensemble of 5 models which together yielded the final labels by the means of majority voting.

The development of the algorithm started at the Department of Computer Science at UIT the Arctic University of Norway. Based on funds from the Norwegian Research Council, a start-up company, Medsensio AS, was established, where the algorithm has been further developed. We evaluate the most recent version.

### Validaton of the algorithm in set A

To reduce the annotation workload, we selected 615 files from the 12090 sound files from the 7^th^ Tromsø study that had not previously been classified. To get a selection with close to equal numbers of files with normal sounds, wheezes, and crackles, we applied a subsetting model to classify the recordings, which was different from the algorithm we were going to validate. It was a single model developed on the same training set from the Tromsø study that was used for the development of the ensemble of models evaluated in this paper, with sparser input features. To further reduce possible bias in the evaluation results favoring the algorithmic solution that could arise by this method of selecting a subset, the following procedure was followed. Firstly, prediction with the use of the subsetting model was performed for all 12090 files. For each class (normal, wheeze, crackle) assigned by the algorithm, 200 files were selected in such a way, that – conditional on the given label being assigned – a quarter of files were randomly selected from the 1^st^ quartile, a quarter from the 2^nd^ etc. and the division into quartiles for a given label was performed based on values of the scores (probabilities) for a given label assigned by the deep learning model. Additionally, 17 files classified as bad quality were incorporated into selection. Within the selection 2 files were predicted by the subsetting model to have both wheezes and crackles present, hence the total of 615 files were selected.

The 615 sound files were classified independently by two medical doctors who were experienced raters (HM and JCAS – coauthors of the paper)). The raters watched spectrograms of the recordings and were blinded for clinical information. Sound files on which the two raters disagreed were annotated again by both together and consensus was reached, which was used as reference (ground truth).

### Validaton of the algorithm in set B

All 120 sound files previously used in the interobserver study^21^ were used. The ratings of the files done by the four participating lung sound researchers were used to establish the ground truth. They had watched spectrograms while listening to the recordings and were blinded for clinical information. The criteria for presence of wheezes and crackles were fulfilled when at least three out of the four expert raters had annotated their presence.

### Statistical analysis

The ability of the algorithm to detect wheezes and crackles was assessed by calculating area under the curve (AUC) of the probability scores against the ground truth through Receiver operating characteristics (ROC) curve analysis. Sensitivity (also called “recall”), specificity, and positive predictive value (PPV, also called “precision”) of the algorithm labels (wheezes and crackles) were calculated). We also calculated the kappa-agreement between these labels and the ground truth. To be able to compare the algorithm with human classificatiuon, we calculated the kappa-agreement between the two raters in sample A, and between each of 24 raters and the ground truth in sample B. We chose to use all these statistical methods to make the results more easy to compare with other studies. SPSS statistical software was used in most of the analyses. The 95% confidence intervals (CI) of sensitivities, specificities, and PPVs were obtained by use of MedCalc® statistical software, the 95% CI of kappa-agreements were calculated by use of Vassarstats®. F1-scores were calculated based on the confusion matrices as 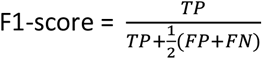, where TP, FP, and FN denote the numbers of true positives, false positives and false negatives, respectively.

This study has been reported according to the STARD guidelines for studies of diagnostic accuracy.^23^

## Results

### Validation in set A

Among the 12090 sound files from which the 615 files in set A were selected, the subsetting model identified wheezes in 1060 (8.8%) and crackles in 592 (4.9%) Among the 615 files selected for validation the algorithm identified 252 with any abnormality and 152 files with wheezes and 105 files with crackles. The corresponding numbers of wheezes and crackles identified by the human raters (ground truth)) were 118 and 126. Among the 17 files rated by the algorithm to be of bad quality, 16 were also found to be of too bad quality to be annotated by the human raters, no lung sound was heard in 13, while three were too noisy to be annotated.

Compared to the ground truth, the algorithms’ prediction scores for wheezes and crackles had an AUC of 0.88 (95% CI 0.84 – 0.92) for wheezes and 0.88 (95% CI 0.84 – 0.92) for crackles (Fig. 2). When the scores were dichotomized, the algorithm detected wheezes with a sensitivity of 81% and a specificity of 89% and crackles with a sensitivity of 67% and a specificity of 97% (Table 1).

**Figure 2.**
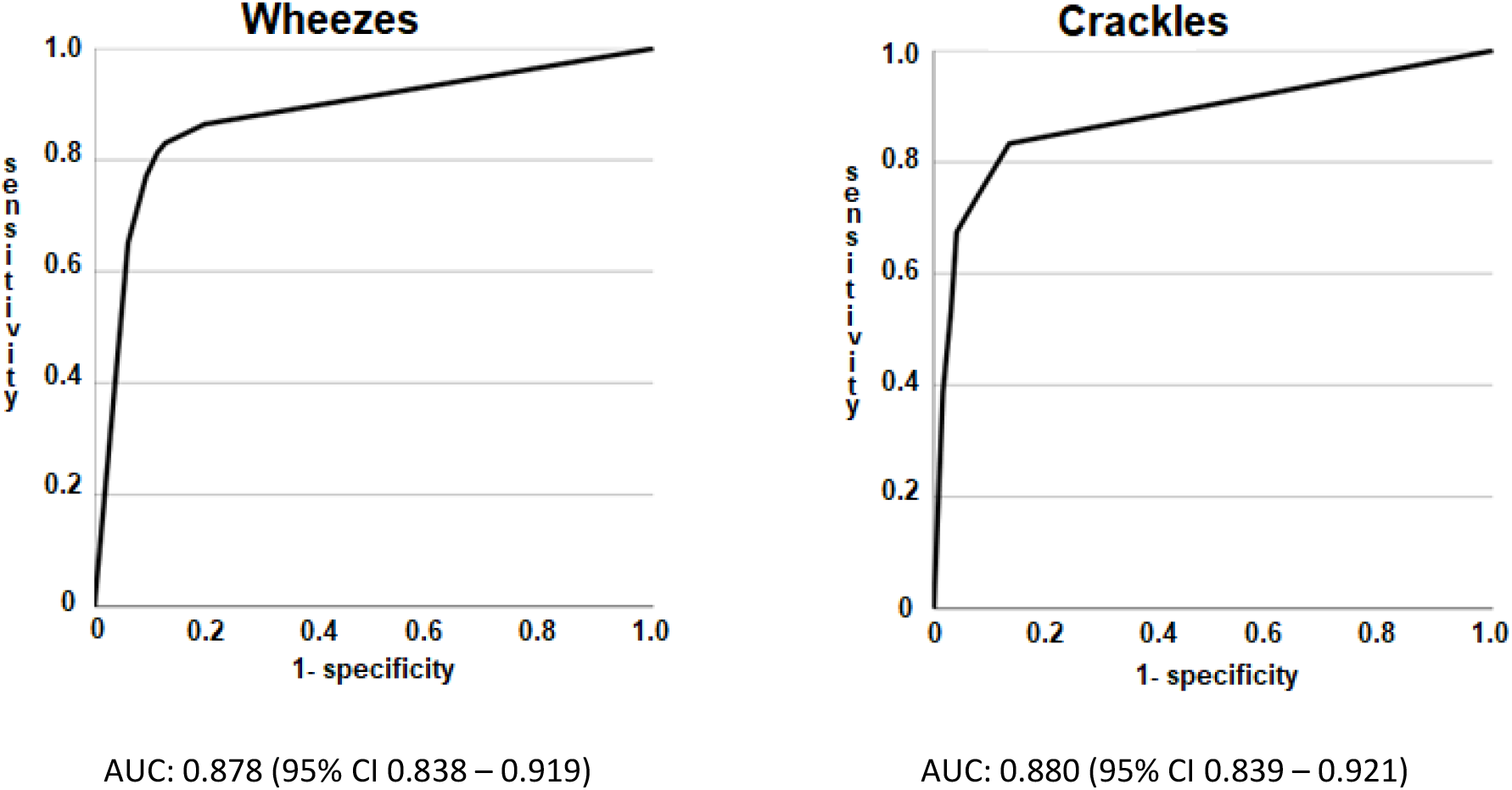
ROC-curves showing the predictive value of the algorithms’ wheeze and crackle scores for ground truth wheezes (n=118) and crackles (n=126) in sample A (615 lung sound recordings)

**Table 1.** 2×2 tables showing sensitivity, specificity, positive predictive value (PPV), and F1-score of the algorithm for detecting wheezes and crackles (ground truth) in lung sound recordings in two separate validation sets, A (615 files from the 7th Tromsø Study) and B (120 files from an interobserver study).

Wheezes
|  |  | Ground truth |  |  |
| --- | --- | --- | --- | --- |
|  |  | Yes | No |  |
| Algorithm | Yes | 96 | 56 | 152 |
|  | No | 22 | 441 | 463 |
|  |  | 118 | 497 | 615 |

Crackles
|  |  | Ground truth |  |  |
| --- | --- | --- | --- | --- |
|  |  | Yes | No |  |
| Algorithm | Yes | 85 | 20 | 105 |
|  | No | 41 | 469 | 510 |
|  |  | 126 | 489 | 615 |

Wheezes
|  |  | Ground truth |  |  |
| --- | --- | --- | --- | --- |
|  |  | Yes | No |  |
| Algorithm | Yes | 8 | 4 | 12 |
|  | No | 0 | 108 | 108 |
|  |  | 8 | 112 | 120 |

Crackles
|  |  | Ground truth |  |  |
| --- | --- | --- | --- | --- |
|  |  | Yes | No |  |
| Algorithm | Yes | 15 | 4 | 150 |
|  | No | 4 | 97 | 446 |
|  |  | 19 | 101 | 120 |

Sensitivity: 79% (95% CI 54% - 94%), Specificity: 96% (95% CI 90% - 99%) PPV: 79% (95% CI 58% - 91%) F1-score 79%

The kappa agreement between the algorithm and the ground truth was 0.631 (95% CI 0.558 – 0.705) for wheezes and 0.68 (95% CI 0.60 – 0.75 for crackles, which was better agreement than with each of the raters (Table 2). For comparison, the corresponding kappa agreements between the two raters were 0.47 and 0.47, respectively.

**Table 2.** Kappa-agreement with 95% confidence interval between algorithm and human raters in validation set A (615 files from the 7th Tromsø Study)

|  | Kappa | (95% CI) |
| --- | --- | --- |
| **Wheezes** |  |  |
| Algorithm versus rater 1 | 0.576 | (0.501 – 0.650) |
| Algorithm versus rater 2 | 0.524 | (0.443 – 0.606) |
| **Algorithm versus ground truth (rater consensus)** | **0.631** | **(0.558 – 0.705)** |
| Rater 1 versus rater 2 | 0.465 | (0.383 – 0.547) |
| **Crackles** |  |  |
| Algorithm versus rater 1 | 0.609 | (0.530 – 0.689) |
| Algorithm versus rater 2 | 0.602 | (0.519 – 0.685) |
| **Algorithm versus ground truth (rater consensus)** | **0.676** | **(0.600 – 0.751)** |
| Rater 1 versus rater 2 | 0.469 | (0.382 – 0.555) |

### Validation in set B

Among the 120 files in sample B, the algorithms’ prediction scores had an AUC of 0.991 (95% CI 0.976 – 1.0) for wheezes and 0.949 (95% CI 0.885 – 1.0) for crackles. Dichotomized, the algorithm detected wheezes with a sensitivity of 100 % and specificity of 96% and crackles with a sensitivity of 79% and specificity of 96% (Table 1), The kappa agreement between the algorithm and the ground truth was 0.783 (95% CI 0.578 – 0.987) for wheezes and 0.749 (95% CI 0.585 – 0.915) for crackles. The kappa agreements between the ground truth and the 24 observers varied between 0.130 and 1.000 for wheezes with a mean of 0.674 and between 0.211 and 0.820 for crackles with a mean of 0.548 (Fig 3).

**Figure 3.**
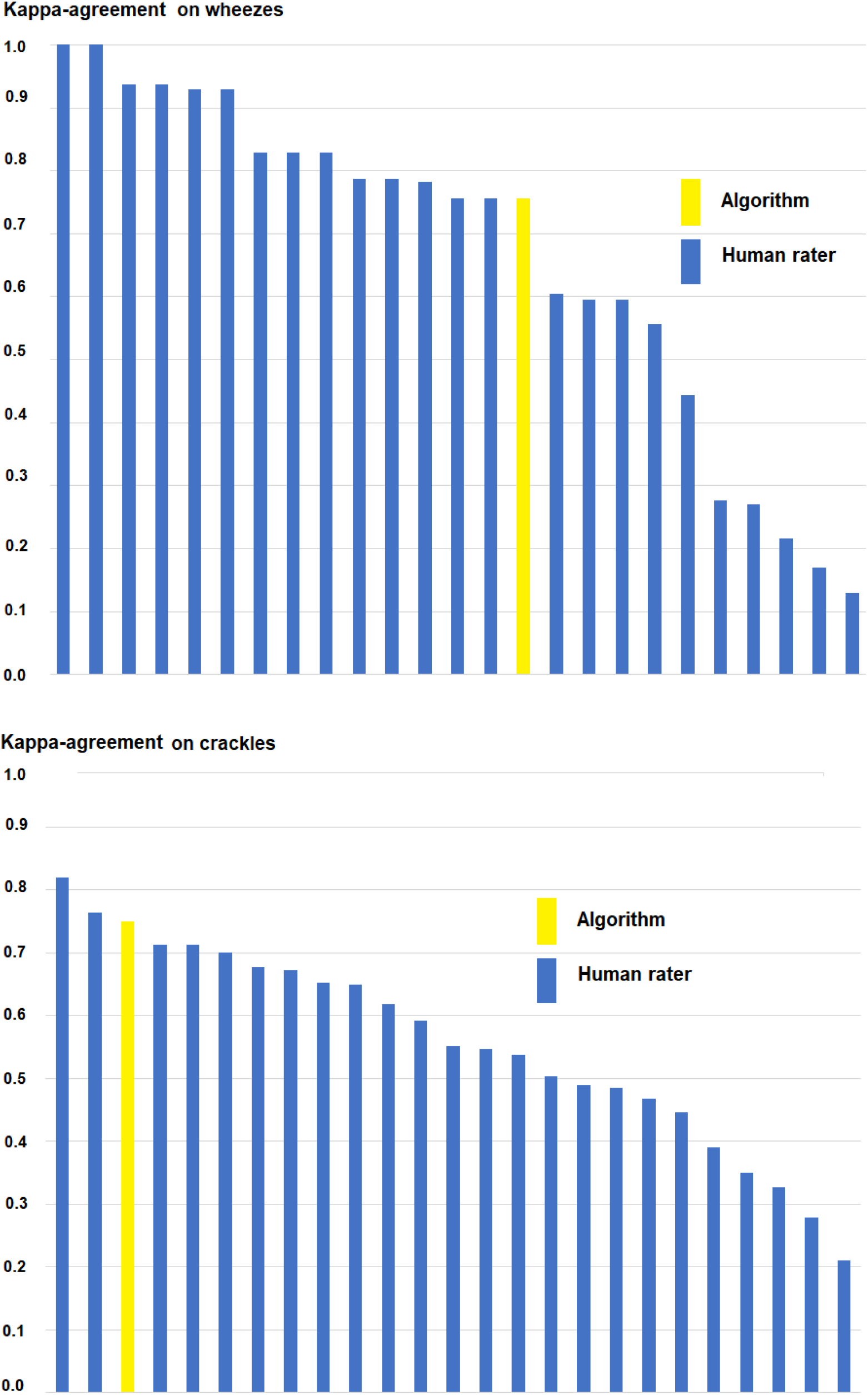
Kappa-agreements between 24 human raters and the ground truth on the presence of wheezes and crackles in sample B (120 lung sound recordings).

## Discussion

The validation of the algorithm showed a promising ability to predict the reference classification. The agreement with reference in set B surpassed the achievement of most of the human raters. Accordingly, lung sound classification done by a computer seems to be more reliable than when done by an average physician. The lung sound files in validation set A were recorded by exactly the same method as the data set that trained the algorithm. The recordings in set B were done with the same electronic stethoscope, but in a different setting. It was still not surprising that the best agreement was found in sample B, since those files were specially selected to be of good quality.

### Comparison with previous studies

We found Kappa agreements in the range 0.6 to 0.8 which is regarded to be substantial ^24^. Studies of agreement between human annotators often show lower values. McCollum and coworkers found kappa-agreements of 0.45 for wheezes and 0.41 for crackles when recorded lung sounds were rated ^25^. Melbye and coworkers found kappa-agreements between experienced observers to be 0.59 for wheezes and 0.62 for crackles^6^. Ferreira-Cardoso and coworkers found a kappa-agreement on any abnormal sound in good-quality recordings of 0.66 ^26^. Three pediatric pulmonologists identified wheezes with a kappa of 0.76 in 55 good-quality recordings from six patients ^27^. In a clinical study of 115 hospitalized patients, similar kappa-agreements as in our study was found for wheezes and fine crackles, but lower values for rhonchi and coarse crackles ^28^.

A few comparable studies have evaluated algorithms for detection of wheezes and crackles. Kim and coworkers found higher concordance between algorithms and the reference classification, but the evaluation was done in sound files that had also been used in the training of the algorithm ^9^. Grzywalski and coworkers found that the algorithm (neural network) detected wheezes and crackles in children with sensitivities ranging from 56% to 88% and with specificities from 79% to 88% ^10^. Compared to our results, these values were generally somewhat lower, but the subdivision of wheezes into wheezes and rhonchi and crackles into fine and coarse may have contributed to lower concordance ^6^. Anyhow, the algorithm obtained higher sensitivity than pediatricians, and similar specificity. Kevat and coworkers found a concordance between the algorithm and the expert classification, also in recordings from children, to be in a similar range as in our study. The sensitivity and specificity for detecting wheezes were 76% - 90% and 95% - 97%, respectively, and for crackles 60%-86% and 96%-99%. They concluded that the algorithm was promising and with at least similar diagnostic accuracy to that of many clinicians ^11^.

### Strengths and limitations

Some strengths are related to the training of the algorithm, others to how the validation has been carried out. The algorithm was based on a large number of recordings, 24090 in total, and rigorous human classifications without access to clinical information. The validation was done in two sets of lung sound recordings, one principally identical to the sample that trained the algorithm and one from a different setting. The latter validation set had been rated by a great number of clinicians, which made it possible to compare the performance of the algorithm with that of relevant human raters. The same stethoscope was used in both validation sets. This might limit the validity of the algorithm when it comes to recordings with different devices. Further, the validated algorithm only outputs whether wheezes and crackles are present or not and it does not give detailed information on quality and timing of the adventitious sounds, which could increase the algorithms’ diagnostic usefulness ^12^. Finally, the algorithm would probably have performed worse if the validation sets had been recorded in more noisy settings ^13^.

## Conclusion

The algorithm validated in this study predicted substantially the presence of wheezes and crackles in lung sound recordings. The algorithm reached higher kappa agreements with the reference classification than what was observed between human raters. However, we cannot be sure that similar good results can be obtained in different settings or when other recording devices are used. These results may enable novel use of lung sounds in clinical and research applications.

## Supporting information

STARD 2015 checklist

## Data Availability

Researchers can apply for access to the The Tromsø Study data at: https://uit.no/research/tromsostudy

## Author Contributions

Conceptualization HM and JR; Sound file annotations HM and JCAS; Algorithm development JR, MP, and LAB; Statistical and formal analysis HM and MP; Writing original draft HM and MP; Writing review and editing HM, JR, MP, LAB and JCAS.

## Data availability

Researchers can apply for access to The Tromsø Study data at: https://uit.no/research/tromsostudy

## Acknowledgments

The Tromsø study was approved by the Norwegian Data Inspectorate and the Regional Ethical Committee of North Norway (REK). Only the sound files and variables classifying the sounds were used, and identification of the participants was not possible. The publication charges for this article have been funded by a grant from the open access publication fund of UiT The Arctic University of Norway.

